# Systematic Review and Meta-analysis on COVID-19 Vaccine Hesitancy

**DOI:** 10.1101/2021.05.15.21257261

**Authors:** Iman Ahmed Fathalla Aboelsaad, Dina Mohamed Hafez, Abdallah Almaghraby, Shaimaa Abdulaziz Abdulmoneim, Samar Ossama El-ganainy, Noha Alaa Hamdy, Ehsan Akram Deghidy, Ahmed El-Sayed Nour El-Deen, Ehab Mohamed Adel Elrewany, Alaa Hamdy Taha Khalil, Karem Mohamed Salem, Samar Galal kabeel, Yasir Ahmed Mohammed Elhadi, Ramy Shaaban, Amr Alnagar, Eman Ahmad Fadel Elsherbeny, Nagwa Ibrahim Elfeshawy, Mohamed Moustafa Tahoun, Ramy Mohamed Ghazy

## Abstract

**Background:** The presented meta-analysis was developed in response to the publication of several studies addressing COVID-19 vaccines hesitancy. We aimed to identify the proportion of vaccine acceptance and rejection, and factors affecting vaccine hesitancy worldwide especially with the fast emergency approval of vaccines.

**Methods:** Online database search was performed, and relevant studies were included with no language restriction. A meta-analysis was conducted using R software to obtain the random effect model of the pooled prevalence of vaccine acceptance and rejection. Egger’s regression test was performed to assess publication bias. Quality assessment was assessed using Newcastle-Ottawa Scale quality assessment tool.

**Results:** Thirty-nine out of 12246 articles met the predefined inclusion criteria. All studies were cross-sectional designs. The pooled proportion of COVID-19 vaccine hesitancy was 17% (95% CI: 14-20) while the pooled proportion of COVID-19 vaccine acceptance was 75% (95% CI: 71-79). The vaccine hesitancy and the vaccine acceptance showed high heterogeneity (I^2^=100%). Case fatality ratio and the number of reported cases had significant effect on the vaccine acceptance as the pooled proportion of vaccine acceptance increased by 39.95% (95% CI: 20.1-59.8) for each 1% increase in case fatality (P<0.0001) and decreased by 0.1% (95% CI: -0.2-0.01) for each 1000 reported case of COVID-19, P= 0.0183).

**Conclusion:** Transparency in reporting the number of newly diagnosed COVID-19 cases and deaths is mandatory as these factors are the main determinants of COVID-19 vaccine acceptance.

## Introduction

The wide use of vaccines has led to decreased mortality and morbidity of different transmissible diseases, this was a crucial factor in elimination of poliomyelitis in the Americas and the worldwide eradication of smallpox (1). Vaccination programs depend on mass vaccination to be able to decrease incidence and prevalence of Vaccine Preventable Diseases (VPD). In addition to the proposed direct protection for vaccinated candidates, wide vaccination scope results in indirect shielding for the overall community by declined conveyance of VPD, thereby dampening the risk of infection for vulnerable individuals in the community (2).

One of the main limiting factors for wide-spread of vaccination programs (especially for newly emerging vaccines) is vaccine hesitancy. The World Health Organization (WHO) named vaccine hesitancy as one of the top ten threats to global health in 2019, calling for research to identify the factors associated with this phenomenon (3). Vaccine hesitancy is defined as a behavior of a delayed vaccine approval or even declined vaccination despite accessible vaccination services (4, 5).

The pandemic COVID-19 caused by the recently discovered coronavirus-2019 (SARS-CoV-2) is strongly influencing the worldwide public health, culture, economy, and human social behavior. Despite all efforts since the beginning of the pandemic there is no approved medicine or treatment to cure COVID-19 till now, whereas vaccine development efforts are taking the highest priority as it can potentially save humanity by inducing immunity against COVID-19 (6).

According to WHO, herd immunity against COVID-19, which is known as population immunity, can be achieved naturally by the exposed people who recovered from the virus by their own protective antibodies or by providing COVID-19 vaccination (7, 8). Herd immunity for COVID-19 can be achieved on 70% of the single vaccinated dose individuals and 90% of the two vaccinated dose individuals (9).

Vaccines typically require years of research and testing before reaching the clinic, but in 2020, scientists were racing against time to produce safe and effective coronavirus vaccines. Currently we have 14 approved vaccines for full use, 6 authorized in early or limited use, 27 vaccines in phase 3 trials, 36 vaccines in phase 2, 48 vaccines in phase 1 and 4 abandoned vaccines after trials. In addition, at least 77 preclinical vaccines are under active investigation in animals (10). Unfortunately, the newly emerging vaccines for COVID-19 are faced nowadays with hesitancy to use in different countries. People showed concerns about both efficacy and possible side effects of these recently approved vaccines. Such hesitancy can have a heavy influence on vaccine delivery and the aimed wide uptake to control the pandemic (11). After the announcement of several pharmaceutical manufactures the production of COVID-19 vaccines, social media started to discuss vaccine content widely across different platforms. The propagated information provides mostly non-factual data and from non-medical individuals (12).

The presented systematic review & meta-analysis was developed in response to the publication of several studies addressing COVID-19 vaccines hesitancy. Identification of independent factors affecting vaccine hesitancy worldwide especially with the fast emergency approval of these vaccines.

## Methods

### Data sources

This meta-analysis was guided by the 2020 Cochrane Handbook of Systematic Review and Meta-Analysis (13), with respect to the preferred reporting items of the systematic review and meta-analysis (PRISMA) checklist (14). Search was conducted for the hesitancy or refusal of COVID-19 vaccination through the published and grey literature using multiple databases; PsycINFO, ScienceDirect, Embase, Scopus, EBSCO, MEDLINE central/PubMed, ProQuest, SciELO, SAGE, Web of science, and Google scholar. Search terms were determined and approved after the consultation of PubMed help desk. The used keywords were added to **Annex 1**.

### Study selection

All studies reporting COVID-19 vaccine hesitancy, were included with no language restriction. Abstract-only papers as proposals, conference, editorials, author responses, reviews, case reports, case series, books and studies with data not accurately or reliably extracted, duplicate, or overlapping data were excluded.

### Data extraction and selection process

**Figure(1)** depicts the PRISMA flow chart for the different steps of the systematic review. All articles were imported into EndNote X7.0.1 to detect and remove the duplicates with two methods: title, author, year, and then manually using title, author, and journal. Title and Abstract screening followed by full text screening were done after the citation’s exportation to an Excel sheet containing the author’s name, publication year, journal, DOI, URL link, and the abstract. Screening was performed independently by 3 reviewers NA Hamdy, EAD fourth reviewer IAA solved any disagreement. The kappa test of agreement between reviewers was 0.89.

**Figure 1:**
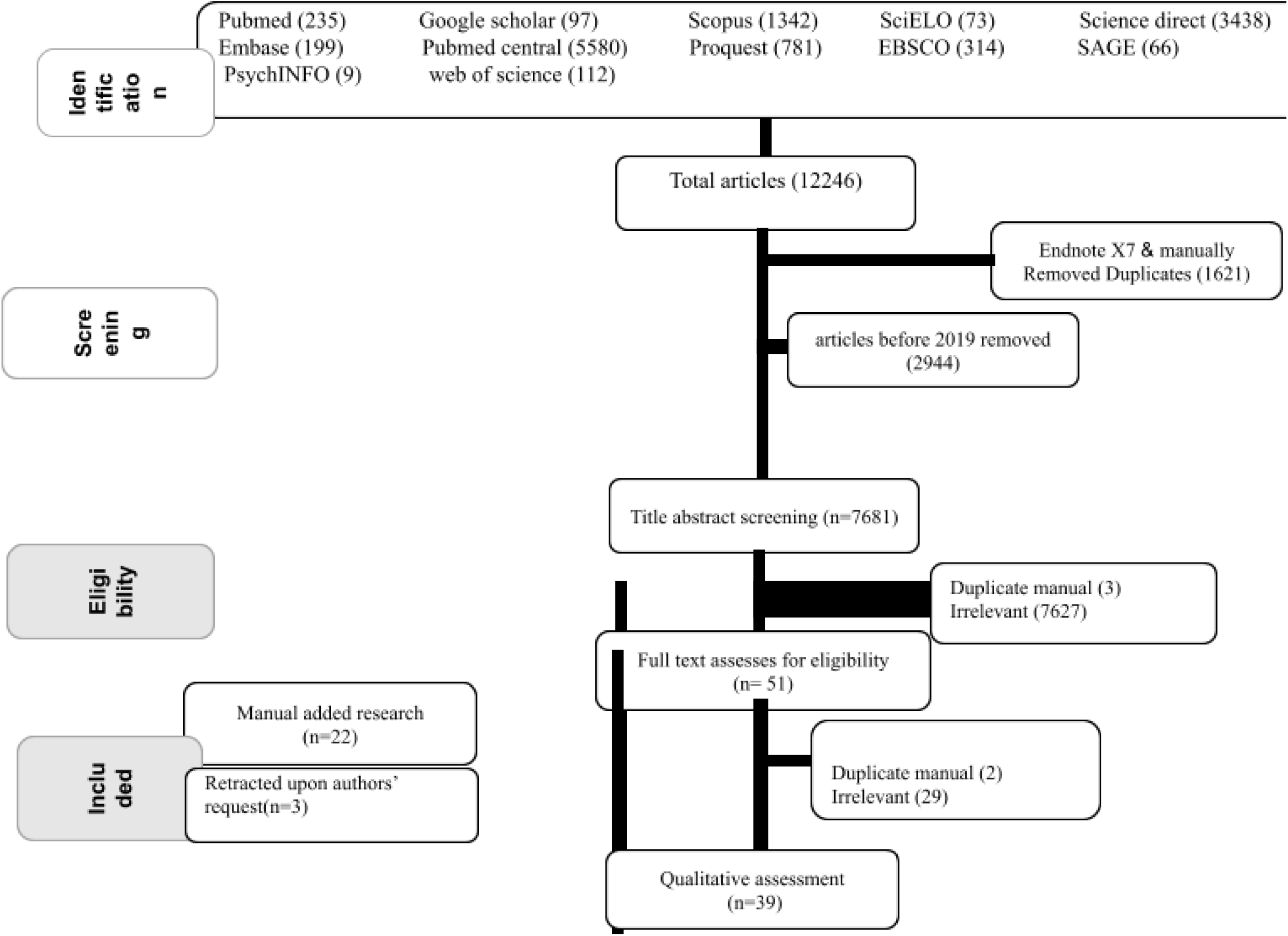
PRISMA flow chart of studies screened and included.

Further manual search for eligible citations was done through careful examination of the references of included studies as well as studies citing the selected articles using PubMed and google scholar. All included articles were extracted to an excel sheet with the following predefined data: year of publication, authors name, country, study design, study setting, study population, sample size, duration of the study, inclusion and exclusion criteria, percent refusal, percent acceptance, cause of refusal and cause of acceptance. Excel sheets are available online for reviewers. At **Annex 2** https://drive.google.com/file/d/12tiK2yW9RGmVnxOTphDKOAuAFyMVub_K/view?usp=sharing

### Investigations of heterogeneity

Cochrane Q test and (I^2^) test was used to assess and measure heterogeneity between studies, considering I^2^ ≥ 75% represents substantial heterogeneity and strength of evidence for heterogeneity is the P-value ≤ 0.05 from the Q test; according to Cochrane Handbook for Systematic Reviews of Interventions (13). Due to substantial heterogeneity, DerSimonian and Laird random-effects models were applied to pool the outcomes.

### Publication bias

Publication biases were assessed by visual inspection of the funnel plot and statistically by Begg’s modified funnel plot and Egger’s regression test (13).

### Quality assessment

Quality assessment (QA) was assessed using Newcastle-Ottawa Scale quality assessment tool customized for cross-sectional studies (15). The assessment was performed by two independent reviewers (DMH, EE) and further checked by two additional reviewers (SO EI-ganainy, AA).

### Statistical analysis and data synthesis

R software was used to perform the meta-analysis and to pool the effect size (proportion); fixed or random effect model were used according to the studies’ consistency. Meta-regression analysis was performed to examine the impact of confounders on the effect of vaccine hesitancy such as age, sex, and country. Results were presented in the Forest plots to visualize the degree of variation between studies. **Leave one-out sensitivity analysis** was conducted to test the effect of each study on the pooled effect to determine the robustness of the obtained outcomes. **Sub-group analysis** was performed to categorize the vaccine hesitancy according to sample size studies.

To investigate the sources of high heterogeneity in the pooled prevalence of vaccine acceptance and hesitancy, meta-regression analysis was performed with different models including the main predictors of vaccine acceptance and hesitancy reported in included studies such as age, sex, educational level and setting. Additionally, number of reported cases, number of reported deaths, case fatality ratio and number of vaccinated people within each country until the end of January 2021 (16, 17), were examined as potential modifiers of vaccine acceptance and hesitancy and included in the meta-regression model.

## Results

### Search results

The flow diagram of the selection process is shown in **figure 1**. From a total of 12246 potentially relevant articles, 1621 duplicate articles and 2944 citations published before 2019 were excluded. A total of 7681 citations were eligible for title screening. Only 51 articles were eligible for full-text screening after removing irrelevant (7627) and duplicate articles (3). In total 34 articles were excluded after full text screening (2 duplicates and 29 irrelevant), 3 were retracted. Another 22 articles were added manually For quantitative assessment, there were 39 eligible articles. The inter-rater agreement for inclusion was κ=0.87 and for the quality assessment was κ=0.91.

**Figure 2** presents the funnel plot of 38 studies reporting the COVID-19 vaccine hesitancy and Eggers’ test [t = -1.215, P-value= 0.232], show absence of asymmetry and disapprove any publication bias. **Figure 3** depicts the studies reporting COVID-19 vaccine acceptance and Eggers’ test [t= -0.64, p-value =0.526].

**Figure 2:**
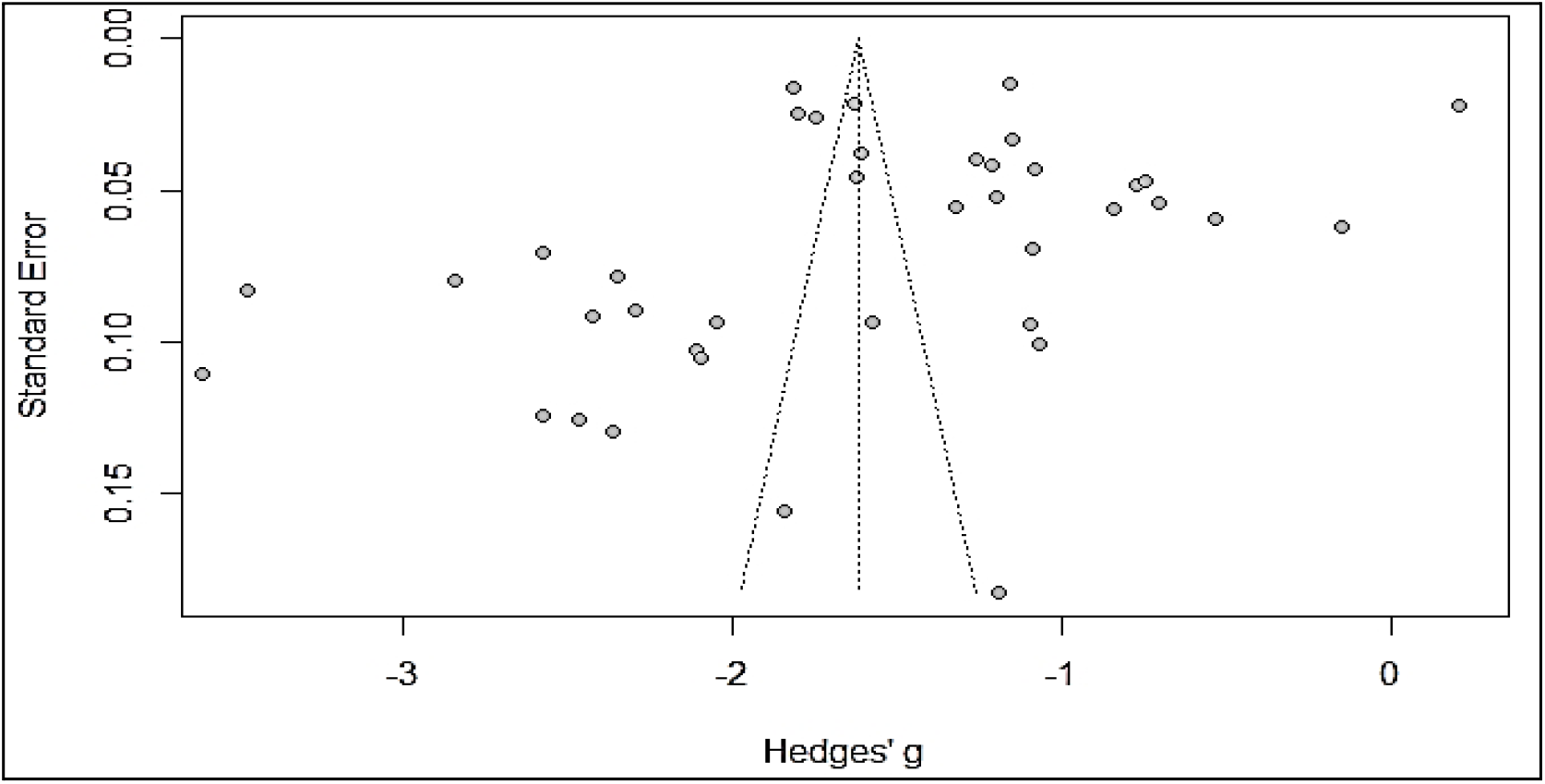
Funnel plot of studies reporting the COVID vaccine hesitancy.

**Figure 3:**
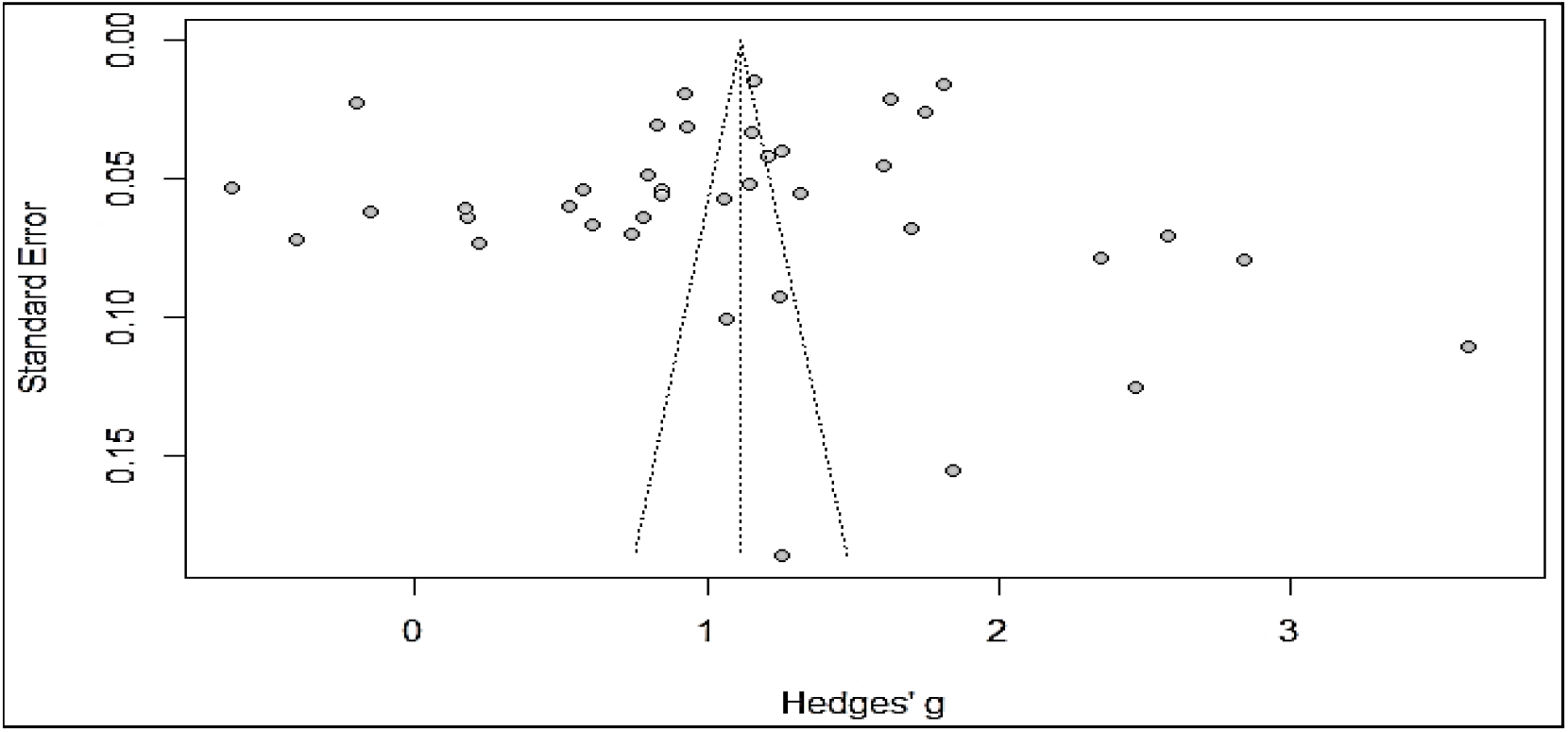
Funnel plot of studies reporting the COVID-19 vaccine acceptance.

**Table 1** shows the main findings of included studies; all the studies were cross-sectional surveys. The total sample size was 173213 ranging from 351 in the study of Sharun *et al*, 2020 (18) reaching 32361 in the study of Paul *et al*, 2021 (19). The highest presentation of female sex was in the study of Kowk, 2021 (20) followed by Wang 2020 (21) while the lowest proportion of females was in the study of Malik *et al* 2020 (22). Age range was 15->85 in the study of Taylor 2020, the mean age of the study participants was the highest in the study of Taylor 2020 (23) (53 years old) and lowest in the study of Al-Mohaithef (24) 31.5 years old. Tools used to assess vaccine hesitancy were online questionnaires either Google forms or Qualtrics forms. Data was collected either <u>through face-to-face interview,</u> online, or both. The quality score of the studies were either very good in 5 studies, good in 20 studies, satisfactory in 12 studies, and unsatisfactory in 5 studies. The main identified predictors of vaccine hesitancy were age, gender, general trust and unknown side effects of the vaccine. The highest vaccine hesitancy were in Wang *et al*, 2020 study (21) (47.8%) and Unroe *et al*, 2020 study (25) (45.1%), while Murphy 2021 *et al*, 2020 study (26) (6%) and Salali and Uysal,, 2020 study (27) (3%) showed the lowest vaccine hesitancy rates.

**Table 1:**
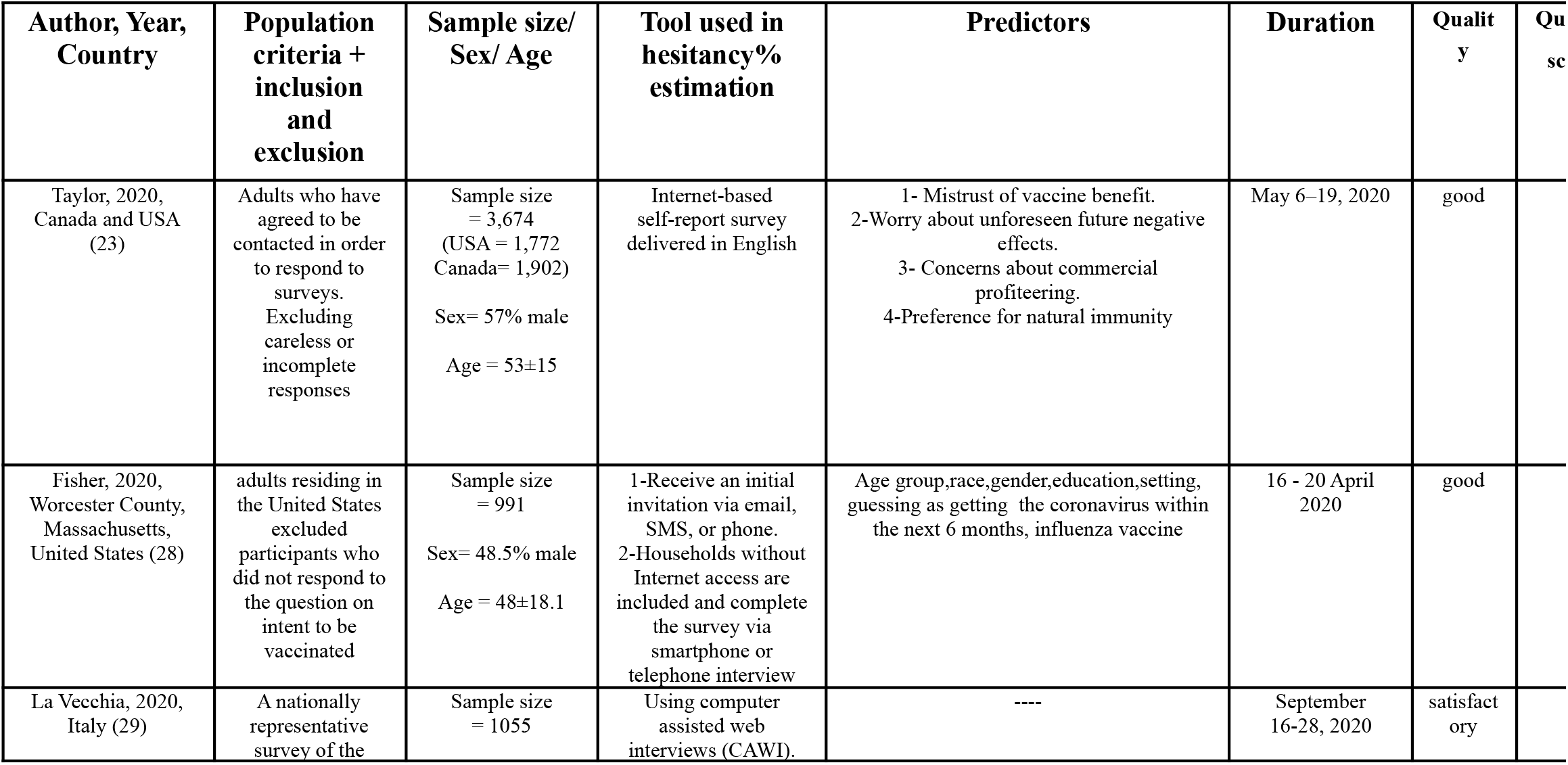
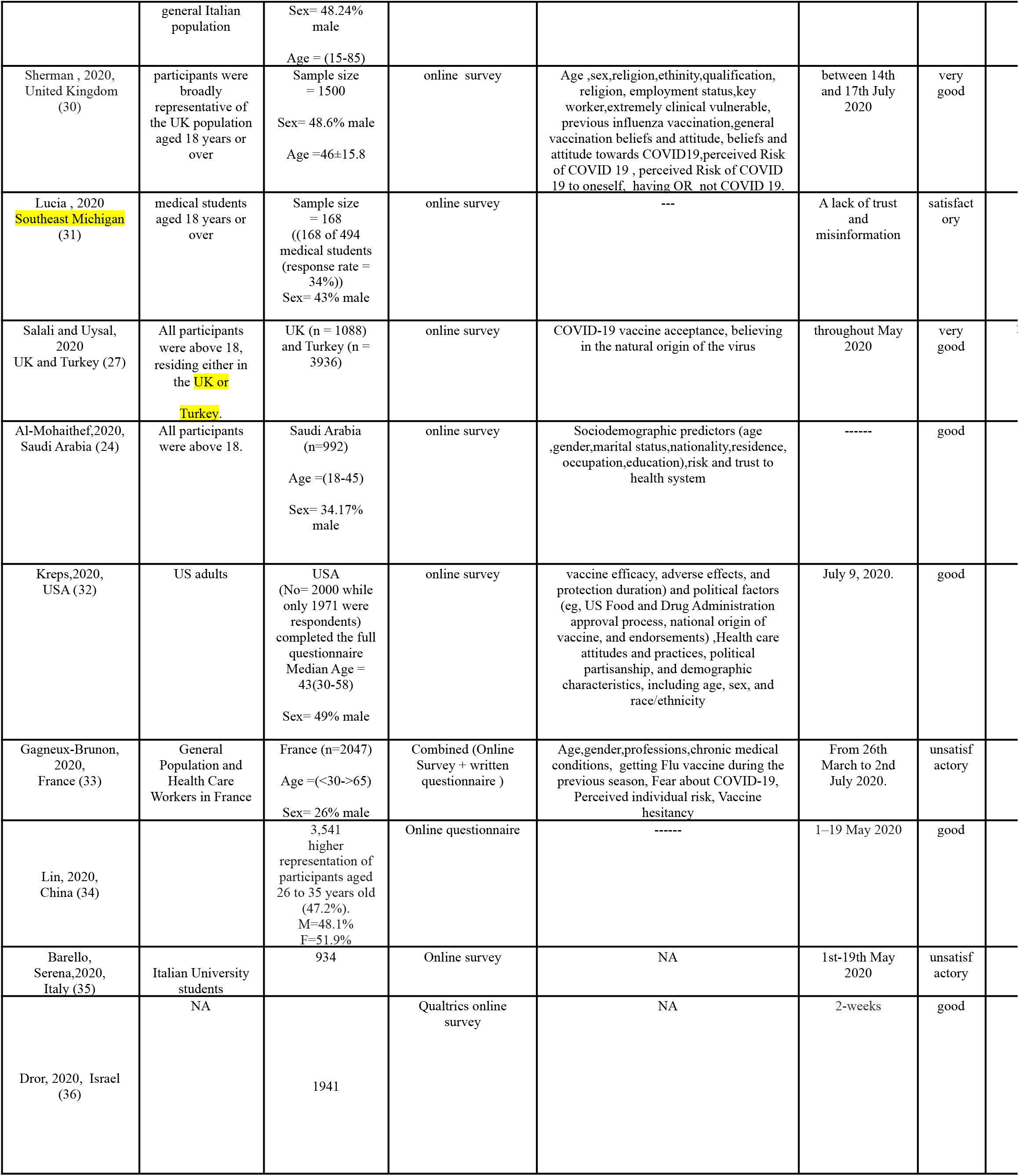
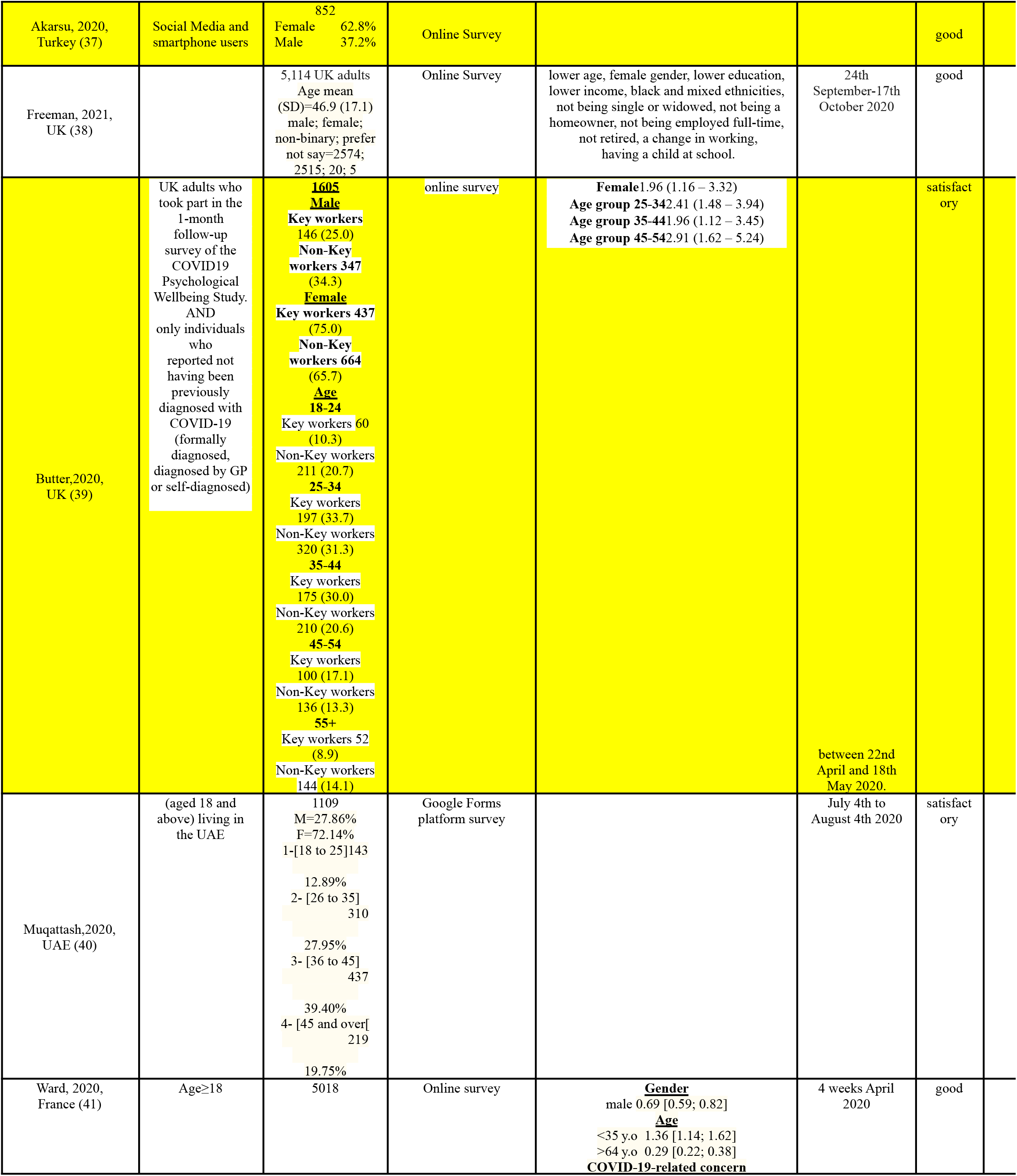
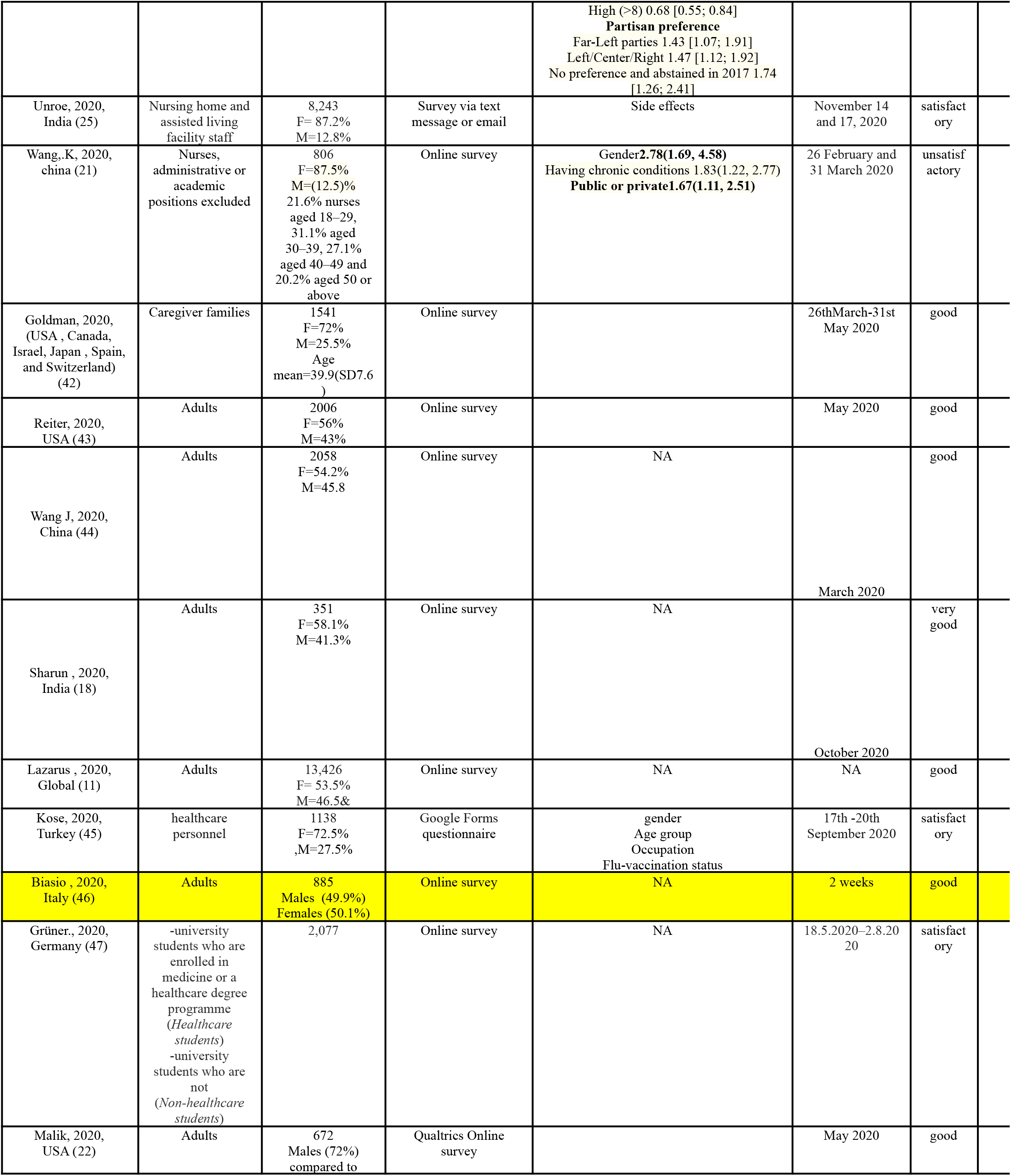
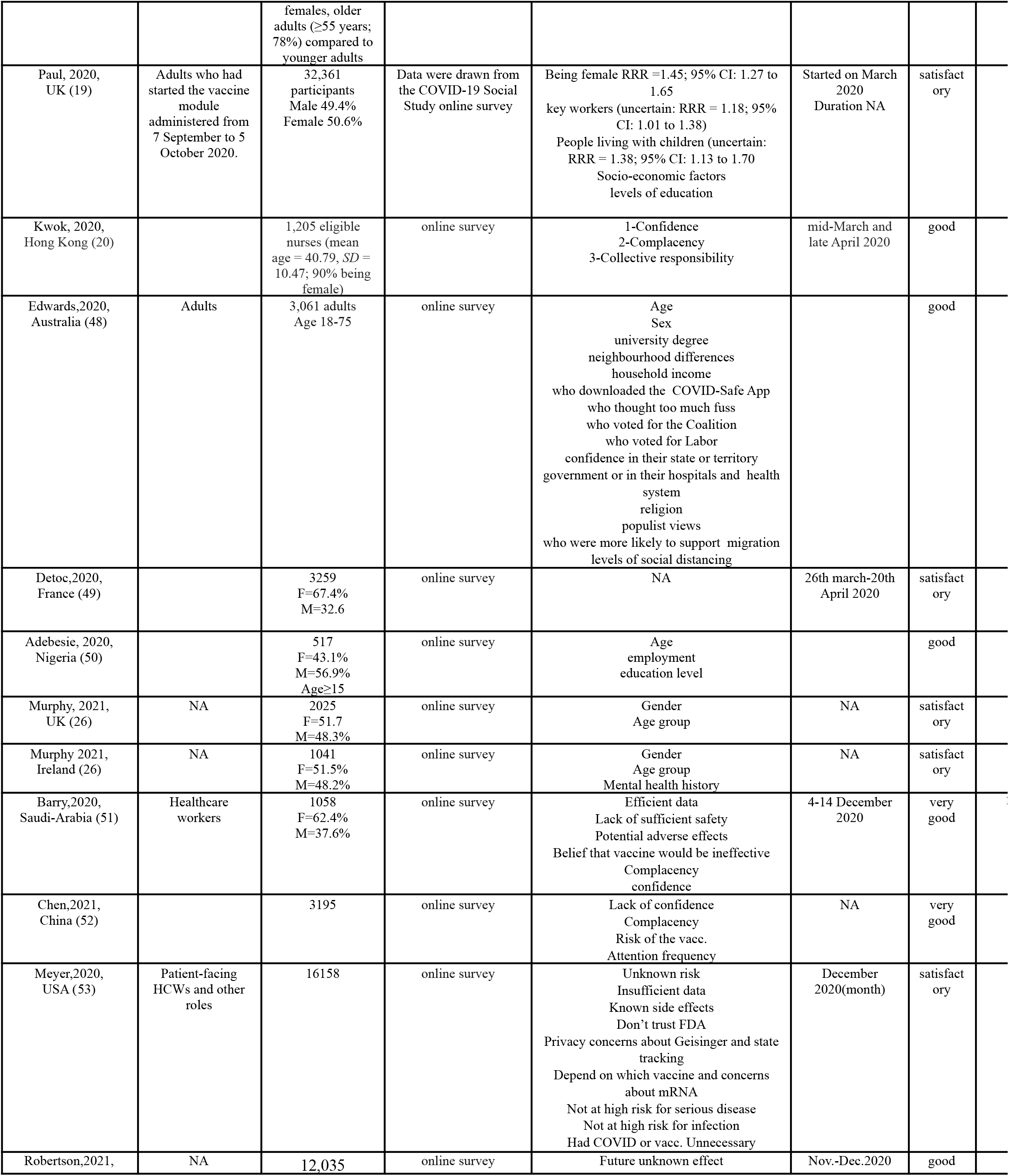
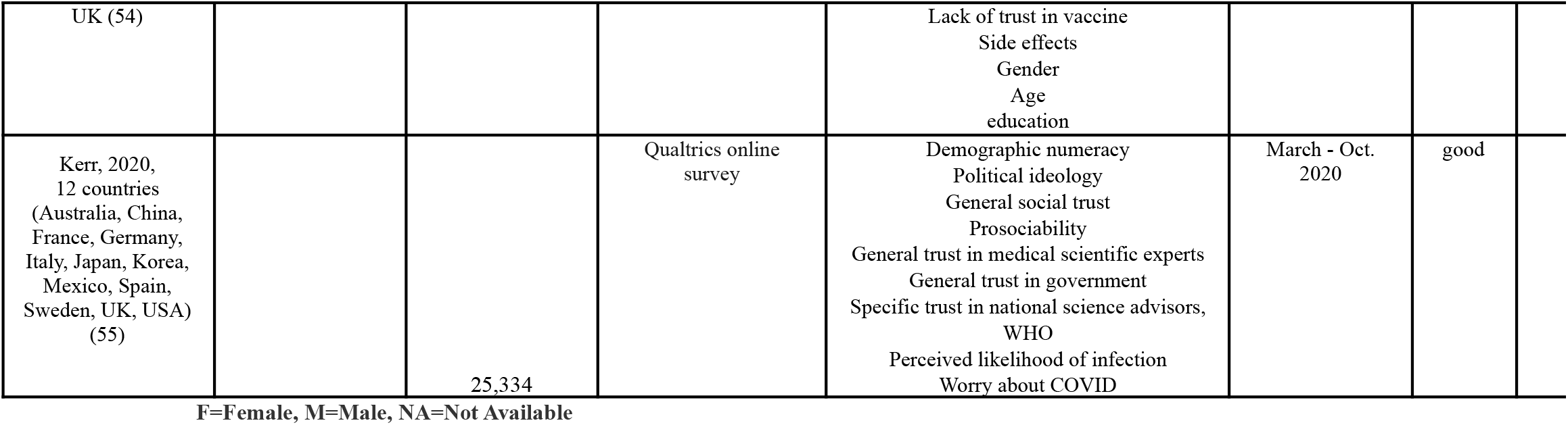
Summary Table of included studies in the meta-analysis.

| Author, Year, Country | Population criteria + inclusion and exclusion | Sample size/ Sex/ Age | Tool used in hesitancy% estimation | Predictors | Duration | Quality | Qu sc |
| --- | --- | --- | --- | --- | --- | --- | --- |
| Taylor, 2020, Canada and USA (23) | Adults who have agreed to be contacted in order to respond to surveys. Excluding careless or incomplete responses | Sample size = 3,674 (USA = 1,772 Canada= 1,902)<br>Sex= 57% male<br>Age = 53±15 | Internet-based self-report survey delivered in English | 1- Mistrust of vaccine benefit.<br>2-Worry about unforeseen future negative effects.<br>3- Concerns about commercial profiteering.<br>4-Preference for natural immunity | May 6–19, 2020 | good |  |
| Fisher, 2020, Worcester County, Massachusetts, United States (28) | adults residing in the United States excluded participants who did not respond to the question on intent to be vaccinated | Sample size = 991<br>Sex= 48.5% male<br>Age = 48±18.1 | 1-Receive an initial invitation via email, SMS, or phone.<br>2-Households without Internet access are included and complete the survey via smartphone or telephone interview | Age group,race,gender,education,setting, guessing as getting the coronavirus within the next 6 months, influenza vaccine | 16 - 20 April 2020 | good |  |
| La Vecchia, 2020, Italy (29) | A nationally representative survey of the | Sample size = 1055 | Using computer assisted web interviews (CAWI). | ---- | September 16-28, 2020 | satisfactory |  |
|  | general Italian population | Sex= 48.24% male<br>Age = (15-85) |  |  |  |  |  |
| Sherman , 2020, United Kingdom (30) | participants were broadly representative of the UK population aged 18 years or over | Sample size = 1500<br>Sex= 48.6% male<br>Age =46±15.8 | online survey | Age ,sex,religion,ethnity,qualification, religion, employment status,key worker,extremely clinical vulnerable, previous influenza vaccination,general vaccination beliefs and attitude, beliefs and attitude towards COVID19,perceived Risk of COVID 19 , perceived Risk of COVID 19 to oneself, having OR not COVID 19. | between 14th and 17th July 2020 | very good |  |
| Lucia , 2020<br>Southeast Michigan (31) | medical students aged 18 years or over | Sample size = 168<br>((168 of 494 medical students (response rate = 34%))<br>Sex= 43% male | online survey | --- | A lack of trust and misinformation | satisfactory |  |
| Salali and Uysal, 2020<br>UK and Turkey (27) | All participants were above 18, residing either in the UK or Turkey. | UK (n = 1088) and Turkey (n = 3936) | online survey | COVID-19 vaccine acceptance, believing in the natural origin of the virus | throughout May 2020 | very good |  |
| Al-Mohaithef,2020, Saudi Arabia (24) | All participants were above 18. | Saudi Arabia (n=992)<br>Age =(18-45)<br>Sex= 34.17% male | online survey | Sociodemographic predictors (age ,gender,marital status,nationality,residence, occupation,education),risk and trust to health system | ----- | good |  |
| Kreps,2020, USA (32) | US adults | USA (No= 2000 while only 1971 were respondents) completed the full questionnaire<br>Median Age = 43(30-58)<br>Sex= 49% male | online survey | vaccine efficacy, adverse effects, and protection duration) and political factors (eg, US Food and Drug Administration approval process, national origin of vaccine, and endorsements) ,Health care attitudes and practices, political partisanship, and demographic characteristics, including age, sex, and race/ethnicity | July 9, 2020. | good |  |
| Gagneux-Brunon, 2020, France (33) | General Population and Health Care Workers in France | France (n=2047)<br>Age =( <30->65)<br>Sex= 26% male | Combined (Online Survey + written questionnaire ) | Age,gender,professions,chronic medical conditions, getting Flu vaccine during the previous season, Fear about COVID-19, Perceived individual risk, Vaccine hesitancy | From 26th March to 2nd July 2020. | unsatisfactory |  |
| Lin, 2020, China (34) |  | 3,541 higher representation of participants aged 26 to 35 years old (47.2%).<br>M=48.1%<br>F=51.9% | Online questionnaire | ----- | 1-19 May 2020 | good |  |
| Barello, Serena,2020, Italy (35) | Italian University students | 934 | Online survey | NA | 1st-19th May 2020 | unsatisfactory |  |
| Dror, 2020, Israel (36) | NA | 1941 | Qualtrics online survey | NA | 2-weeks | good |  |
| Akarsu, 2020, Turkey (37) | Social Media and smartphone users | 852<br>Female 62.8%<br>Male 37.2% | Online Survey |  |  | good |  |
| Freeman, 2021, UK (38) |  | 5,114 UK adults<br>Age mean (SD)=46.9 (17.1)<br>male; female; non-binary; prefer not say=2574; 2515; 20; 5 | Online Survey | lower age, female gender, lower education, lower income, black and mixed ethnicities, not being single or widowed, not being a homeowner, not being employed full-time, not retired, a change in working, having a child at school. | 24th September-17th October 2020 | good |  |
| Butter, 2020, UK (39) | UK adults who took part in the 1-month follow-up survey of the COVID19 Psychological Wellbeing Study. AND only individuals who reported not having been previously diagnosed with COVID-19 (formally diagnosed, diagnosed by GP or self-diagnosed) | **1605**<br>**Male**<br>**Key workers 146 (25.0)**<br>**Non-Key workers 347 (34.3)**<br>**Female**<br>**Key workers 437 (75.0)**<br>**Non-Key workers 664 (65.7)**<br>**Age 18-24**<br>**Key workers 60 (10.3)**<br>**Non-Key workers 211 (20.7)**<br>**25-34**<br>**Key workers 197 (33.7)**<br>**Non-Key workers 320 (31.3)**<br>**35-44**<br>**Key workers 175 (30.0)**<br>**Non-Key workers 210 (20.6)**<br>**45-54**<br>**Key workers 100 (17.1)**<br>**Non-Key workers 136 (13.3)**<br>**55+**<br>**Key workers 52 (8.9)**<br>**Non-Key workers 144 (14.1)** | online survey | **Female 1.96 (1.16 – 3.32)**<br>**Age group 25-34 2.41 (1.48 – 3.94)**<br>**Age group 35-44 1.96 (1.12 – 3.45)**<br>**Age group 45-54 2.91 (1.62 – 5.24)** | between 22nd April and 18th May 2020. | satisfactory |  |
| Muqattash, 2020, UAE (40) | (aged 18 and above) living in the UAE | 1109<br>M=27.86%<br>F=72.14%<br>1- [18 to 25] 143<br>12.89%<br>2- [26 to 35] 310<br>27.95%<br>3- [36 to 45] 437<br>39.40%<br>4- [45 and over] 219<br>19.75% | Google Forms platform survey |  | July 4th to August 4th 2020 | satisfactory |  |
| Ward, 2020, France (41) | Age ≥ 18 | 5018 | Online survey | **Gender**<br>male 0.69 [0.59; 0.82]<br>**Age**<br><35 y.o 1.36 [1.14; 1.62]<br>>64 y.o 0.29 [0.22; 0.38]<br>**COVID-19-related concern** | 4 weeks April 2020 | good |  |
|  |  |  |  | High (>8) 0.68 [0.55; 0.84]<br>**Partisan preference**<br>Far-Left parties 1.43 [1.07; 1.91]<br>Left/Center/Right 1.47 [1.12; 1.92]<br>No preference and abstained in 2017 1.74 [1.26; 2.41] |  |  |  |
| Unroe, 2020, India (25) | Nursing home and assisted living facility staff | 8,243<br>F= 87.2%<br>M=12.8% | Survey via text message or email | Side effects | November 14 and 17, 2020 | satisfactory |  |
| Wang, K, 2020, china (21) | Nurses, administrative or academic positions excluded | 806<br>F=87.5%<br>M=(12.5)%<br>21.6% nurses aged 18–29, 31.1% aged 30–39, 27.1% aged 40–49 and 20.2% aged 50 or above | Online survey | Gender**2.78(1.69, 4.58)**<br>Having chronic conditions 1.83(1.22, 2.77)<br>**Public or private1.67(1.11, 2.51)** | 26 February and 31 March 2020 | unsatisfactory |  |
| Goldman, 2020, (USA , Canada, Israel, Japan , Spain, and Switzerland) (42) | Caregiver families | 1541<br>F=72%<br>M=25.5%<br>Age mean=39.9(SD7.6) | Online survey |  | 26thMarch-31st May 2020 | good |  |
| Reiter, 2020, USA (43) | Adults | 2006<br>F=56%<br>M=43% | Online survey |  | May 2020 | good |  |
| Wang J, 2020, China (44) | Adults | 2058<br>F=54.2%<br>M=45.8 | Online survey | NA | March 2020 | good |  |
| Sharun , 2020, India (18) | Adults | 351<br>F=58.1%<br>M=41.3% | Online survey | NA | October 2020 | very good |  |
| Lazarus , 2020, Global (11) | Adults | 13,426<br>F= 53.5%<br>M=46.5& | Online survey | NA | NA | good |  |
| Kose, 2020, Turkey (45) | healthcare personnel | 1138<br>F=72.5%<br>M=27.5% | Google Forms questionnaire | gender<br>Age group<br>Occupation<br>Flu-vaccination status | 17th -20th September 2020 | satisfactory |  |
| Biasio , 2020, Italy (46) | Adults | 885<br>Males (49.9%)<br>Females (50.1%) | Online survey | NA | 2 weeks | good |  |
| Grüner., 2020, Germany (47) | -university students who are enrolled in medicine or a healthcare degree programme (*Healthcare students*)<br>-university students who are not (*Non-healthcare students*) | 2,077 | Online survey | NA | 18.5.2020–2.8.2020 | satisfactory |  |
| Malik, 2020, USA (22) | Adults | 672<br>Males (72%)<br>compared to females, older adults ( $\geq 55$ years; 78%) compared to younger adults | Qualtrics Online survey | | May 2020 | good | |
| Paul, 2020, UK (19) | Adults who had started the vaccine module administered from 7 September to 5 October 2020. | 32,361 participants<br>Male 49.4%<br>Female 50.6% | Data were drawn from the COVID-19 Social Study online survey | Being female RRR = 1.45; 95% CI: 1.27 to 1.65<br>key workers (uncertain: RRR = 1.18; 95% CI: 1.01 to 1.38)<br>People living with children (uncertain: RRR = 1.38; 95% CI: 1.13 to 1.70)<br>Socio-economic factors<br>levels of education | Started on March 2020<br>Duration NA | satisfactory |  |
| Kwok, 2020, Hong Kong (20) |  | 1,205 eligible nurses (mean age = 40.79, *SD* = 10.47; 90% being female) | online survey | 1-Confidence<br>2-Complacency<br>3-Collective responsibility | mid-March and late April 2020 | good |  |
| Edwards, 2020, Australia (48) | Adults | 3,061 adults<br>Age 18-75 | online survey | Age<br>Sex<br>university degree<br>neighbourhood differences<br>household income<br>who downloaded the COVID-Safe App<br>who thought too much fuss<br>who voted for the Coalition<br>who voted for Labor<br>confidence in their state or territory government or in their hospitals and health system<br>religion<br>populist views<br>who were more likely to support migration<br>levels of social distancing |  | good |  |
| Detoc, 2020, France (49) |  | 3259<br>F=67.4%<br>M=32.6 | online survey | NA | 26th march-20th April 2020 | satisfactory |  |
| Adebesie, 2020, Nigeria (50) | | 517<br>F=43.1%<br>M=56.9%<br>Age $\geq 15$ | online survey | Age<br>employment<br>education level | | good | |
| Murphy, 2021, UK (26) | NA | 2025<br>F=51.7<br>M=48.3% | online survey | Gender<br>Age group | NA | satisfactory |  |
| Murphy 2021, Ireland (26) | NA | 1041<br>F=51.5%<br>M=48.2% | online survey | Gender<br>Age group<br>Mental health history | NA | satisfactory |  |
| Barry, 2020, Saudi-Arabia (51) | Healthcare workers | 1058<br>F=62.4%<br>M=37.6% | online survey | Efficient data<br>Lack of sufficient safety<br>Potential adverse effects<br>Belief that vaccine would be ineffective<br>Complacency<br>confidence | 4-14 December 2020 | very good |  |
| Chen, 2021, China (52) |  | 3195 | online survey | Lack of confidence<br>Complacency<br>Risk of the vacc.<br>Attention frequency | NA | very good |  |
| Meyer, 2020, USA (53) | Patient-facing HCWs and other roles | 16158 | online survey | Unknown risk<br>Insufficient data<br>Known side effects<br>Don't trust FDA<br>Privacy concerns about Geisinger and state tracking<br>Depend on which vaccine and concerns about mRNA<br>Not at high risk for serious disease<br>Not at high risk for infection<br>Had COVID or vacc. Unnecessary | December 2020(month) | satisfactory |  |
| Robertson, 2021, | NA | 12,035 | online survey | Future unknown effect | Nov.-Dec.2020 | good |  |
| UK (54) |  |  |  | Lack of trust in vaccine<br>Side effects<br>Gender<br>Age<br>education |  |  |  |
| Kerr, 2020,<br>12 countries<br>(Australia, China,<br>France, Germany,<br>Italy, Japan, Korea,<br>Mexico, Spain,<br>Sweden, UK, USA)<br>(55) |  | 25,334 | Qualtrics online<br>survey | Demographic numeracy<br>Political ideology<br>General social trust<br>Prosociality<br>General trust in medical scientific experts<br>General trust in government<br>Specific trust in national science advisors,<br>WHO<br>Perceived likelihood of infection<br>Worry about COVID | March - Oct.<br>2020 | good |  |

F=Female, M=Male, NA=Not Available

## Predictors of COVID-19 vaccine acceptance and hesitancy

Multiple factors were associated with vaccine hesitancy Table(1). Previously receiving influenza vaccine is the main factor that determines the acceptance of COVID-19 vaccine. Individuals reporting intake of influenza vaccine were more likely to accept COVID-19 vaccine than those who did not receive it previously (21, 28, 33).Some socio-demographic characteristics were considered to influence the acceptance of the vaccine. Being young was associated with no or not sure response towards the intake of COVID-19 vaccine (28, 38, 41), while older individuals were more likely to accept the vaccine intake (24, 33). Regarding the gender, males were more likely to accept the vaccine rather than females (21, 33, 38, 45).. Low education levels and income, being not employed in a full time job or retired were associated with refusal of the vaccine(19, 28, 38, 41), while those with professional private work were more likely to accept the vaccine (21). The marital status also affects the response to vaccine acceptance, being single or widowed were associated with hesitancy (38), while married individuals were more likely to accept the vaccine (24). Racial and ethnic groups were noticed to affect the acceptance of vaccine. Black race and mixed ethnicity were associated with hesitancy towards the vaccine (28, 38).Other factors that increase the acceptance towards the vaccine is the presence of trusted health systems (24), the fear from getting infected with the virus (33) and having chronic diseases (21). While factors that increase the refusal of the vaccine involve the suspicion from its efficacy and effectiveness (21), individuals may think about the side effects and do not believe that the vaccine will work, or they trust their immune system and are not afraid of getting sick (45).

### Pooled proportion of COVID-19 vaccine hesitancy and acceptance

Using the random effect model, (**figure 5**) the pooled proportion of COVID-19 vaccine hesitancy among 173213 participants recruited from 39 studies was 17% (95% CI: 14-20). Vaccine hesitancy ranged from 55% (95% CI: 85-87) in the study of Unroe, 2020 (25) to 3% (95% CI: 3-4) in the study of Salali, 2020 (27) and 3% (95% CI: 2-3) in Chen, 2021 (52), with high heterogeneity (I^2^ = 100%). To identify the cause of such heterogeneity, researchers conducted Leave one out sensitivity analysis, sub-group analysis, or meta-regression, however, these analyses failed to explain this heterogeneity. On the other hand, the pooled proportion of COVID-19 vaccine acceptance (**figure 6**) was 75% (95%CI: 71-79). The vaccine acceptance was the highest in Chen, 2021 (97, 95% CI =97-98) and the lowest in Goldman, 2020 (35, 95% CI =32-37). Similar to the vaccine hesitancy, the vaccine acceptance showed high heterogeneity (I^2^=100%). However, meta-regression revealed that case fatality, sample size, the number of reported cases per country and the type of study setting explained 38.52% of the model heterogeneity (p-value <0.0001), the estimated amount of residual heterogeneity (*T*^2^) was 0.3201 (SE = 0.1350). However, only case fatality and the number of reported cases had a significant effect on vaccine acceptance. The pooled proportion of vaccine acceptance increased by 39.95% (95% CI = 20.1-59.8) for each 1% increase in case fatality (*p*<0.0001) and decreased by 0. 1% (95% CI = -0.2 to -0.01) for each 1000 reported case of COVID-19 (*p* = 0.0183).Figure (4) shows the results of the meta-regression models between the case fatality (%) and the proportion of vaccine hesitancy and vaccine acceptance, respectively by type of setting and study sample size.

**Figure (4).**
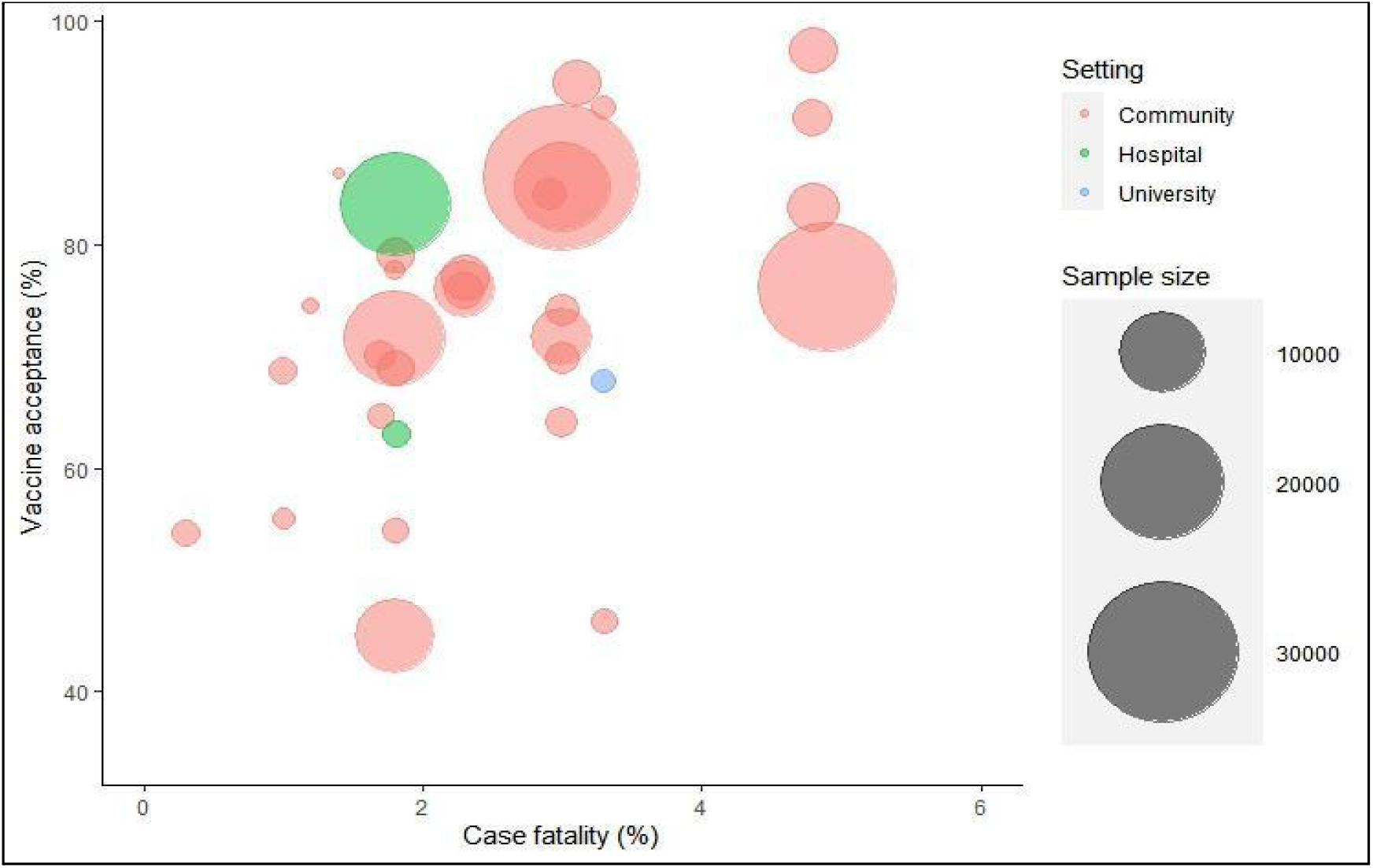
the relation between vaccine acceptance (%) and case fatality (%) by sample size and study setting.

**Figure (5).**
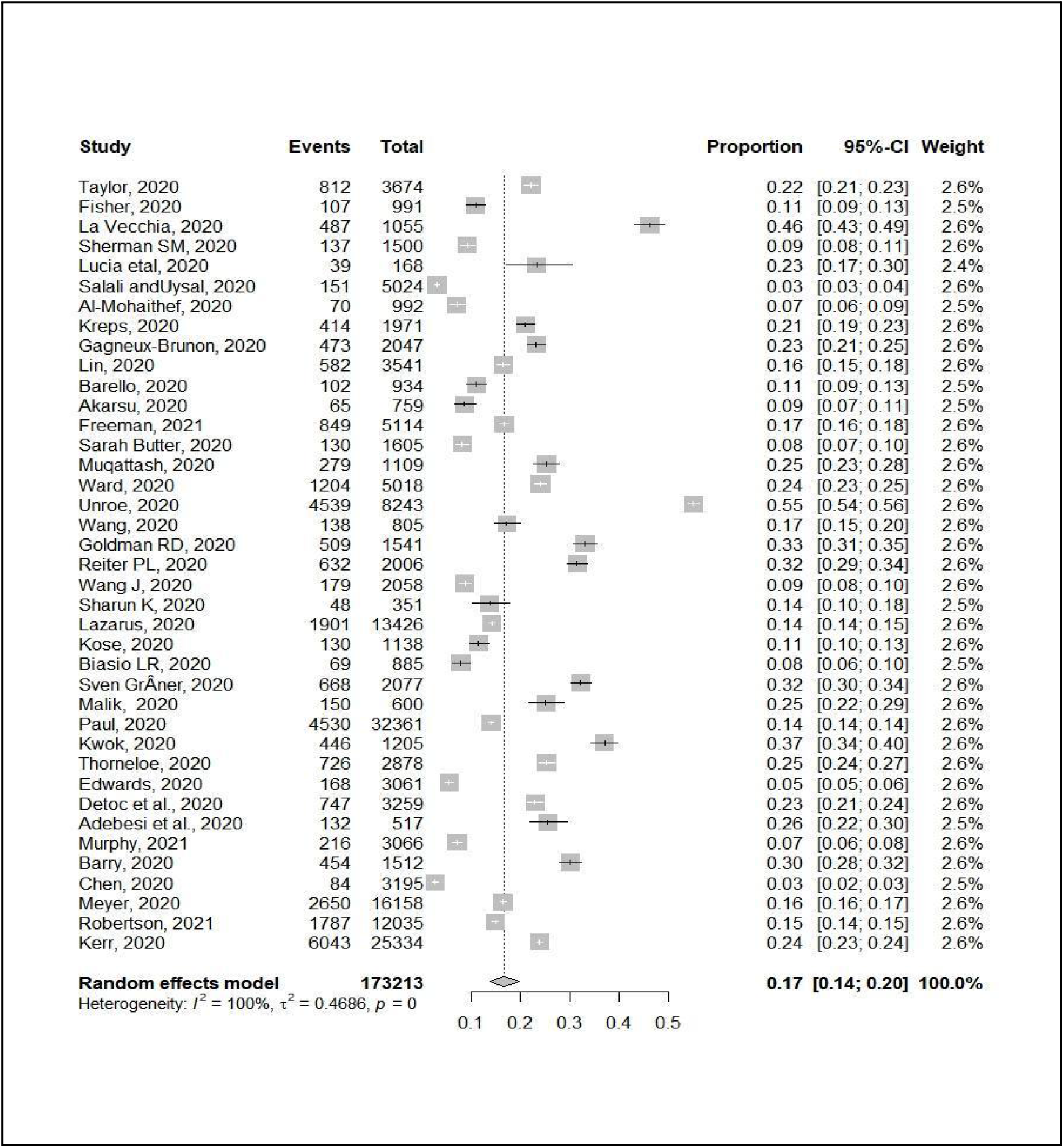
Forest plot of pooled prevalence of vaccine hesitancy.

**Figure (6).**
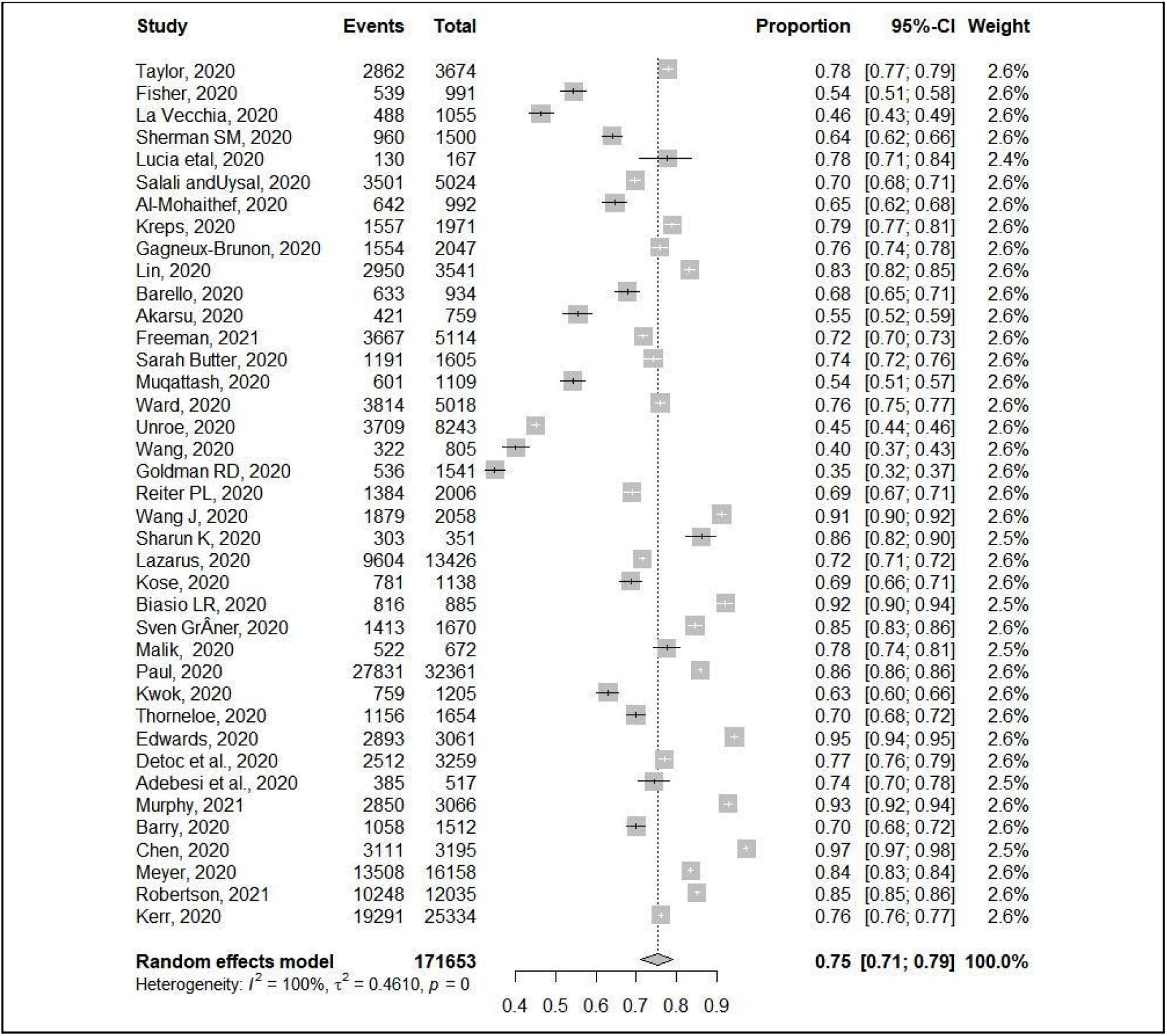
Forest plot of the pooled vaccine acceptance.

## Discussion

The vaccine for COVID-19 availability is a critical step to face the COVID-19 pandemic. But vaccine hesitancy represents a great threat to global health during this pandemic and limits the power of health systems to control the COVID-19 pandemic. Hence, estimating the COVID-19 vaccine hesitancy represents a tool to design an action plan to improve the vaccine acceptance.

In this meta-analysis, there was large variability between the studies discussing COVID-19 hesitancy in terms of vaccine acceptance. We aimed to determine the proportion of the population who are rejecting and accepting COVID-19 vaccine worldwide. We included 39 cross-sectional surveys conducted in 21 countries requiting 173213 participants. The quality of studies ranged from unsatisfactory, to very good. Our meta-analysis revealed that the pooled proportion of COVID-19 vaccine hesitancy was 17% (95% CI: 14-20) while the pooled proportion of COVID-19 vaccine acceptance was 75% (95% CI: 71-79). The main reported determinant of vaccine acceptance was case fatality and number of reported cases. Time effect was not associated with vaccine acceptance.

In the same line, a rapid systematic review and meta-analysis on COVID 19 vaccine hesitancy was conducted by Ronbinson *et al*, (61) to estimate the proportion of individuals refusing COVID-19 vaccine in 13 countries among 58,656 individuals. They reported that about 20% of the participants refused COVID-19 vaccine. They observed that differences across countries were very substantial and resulted in a heterogeneity above 90%. Furthermore, they declared that the trend of rejection increased with time. The main determinants of COVID19 vaccine rejection were being female, of low educational level, or belonging to minor ethnicity.

Another review conducted by Lin *et al*, (62) compared trends in vaccination receptivity over time across US and international polls. The data sources included academic literature, news and official reports published by 20 October 2020. A total of 126 studies and surveys were included. The authors reported that there was a noticeable decline in vaccine acceptance (from >70% in March to <50% in October) with demographic, socioeconomic, and partisan divides observed. Perceived risk, concerns over vaccine safety and effectiveness, doctors’ recommendations, and inoculation history were common factors. Impacts of regional infection rates, gender, and personal COVID-19 experience were inconclusive. Unique COVID-19 factors included political party orientation, doubts toward expedited development/approval process, and perceived political interference. Many receptive participants preferred to wait until others have taken the vaccine; mandates could increase resistance.

We speculate that the difference in vaccine acceptance may be affected by vaccine efficacy and side effects. Vaccines’ side effects range between local to systemic, and short to long term events. The reported common side effects are generally mild to moderate and last for a few days. These include injection site pain, fatigue, rigors, fever, muscle and joints pains. Less commonly, a vaccine recipient may develop allergic reaction or anaphylaxis, and neurological side effects; however they are rarely reported (63). There is a rising concern particularly related to reported thrombo-embolic events, particularly after administration of AstraZeneca vaccine in Europe, but the European Medicines Agency concluded that the benefits of the vaccine overweighs the potential risk of this rare side effect (64). In this context, Kaplan *et al*, (65) underlined that vaccine acceptance improved when vaccine efficacy exceeds 70%. Moreover, they addressed that minor side effects, such as a sore arm or fever lasting for a day did not affect vaccine acceptance, while major side effects in 1/100000 greatly affected vaccine acceptance. These side effects may vary according to the type of vaccine used in each country. Emerging evidence suggests that both exposure to misinformation about COVID-19 and public concerns over the safety of vaccines may be contributing to the observed decline in intentions to be vaccinated, and this highlights the need for measures to address public acceptability, trust and concern over the safety and benefit of approved vaccines (66, 67). This finding highlights the power of social media. Some studies emerged in the last months discussing the vaccine confidence in several populations, especially in countries with high burden of diseases like Pakistan (68). The role of recent misinformation was evident in the study of Loomba *et al*, (Measuring the impact of COVID-19 vaccine misinformation on vaccination intent in the UK and USA), it induced a decline in intent of 6.2 percentage points in the UK and 6.4 percentage points in the USA, among those who stated that they would definitely accept a vaccine. From another perspective, other studies analyzed attitudes toward COVID vaccination like the impact of education, whether medical or nonmedical students, on their decision (35).With the development of multiple effective vaccines, Immunization programs are only successful when there are high rates of acceptance and coverage (69). To accomplish this, it is critical to understand vaccine-acceptance messaging to effectively control the pandemic and prevent thousands of additional deaths (70).Individuals commonly considered COVID-19 to be a very severe disease, although they expected to experience less severe symptoms themselves. Individuals also worried more about transmitting the disease to others than about falling ill personally (71).

The strongest predictor of intentions to accept a COVID-19 vaccine recommended by authorities was the degree to which respondents trusted the vaccine to be safe. Perceived vaccine safety explained 52% of the variance in intentions to vaccinate (72).The study of Malik *et al*. shows that COVID-19 vaccine acceptance can be predicted with relatively high accuracy by readily available demographic characteristics. Since the beginning of the COVID-19 pandemic in the United States, it has been clear that low-income and communities of color are at higher risk for infection and death from COVID-19 (22).

## Strengths and limitations

One of the main strength points in this study is the search strategy, we searched 12 different databases. Each citation was screened by two reviewers and disagreement was solved by a senior author. The same was done for quality assessment to ensure robust evidence. A large proportion of the included studies used quota (as opposed to probability-based sampling) and were pre-prints yet to be peer reviewed (as opposed to published journal articles). However, the type of sampling method used (quota vs. probability) had minimal impact on intentions estimates and that studies reported in pre-prints produced similar effect estimates as peer-reviewed journals. One of the main limitations was different tools used to assess vaccine acceptance in addition, the data collected either through face-to-face interview or through online data collection tools. We think that this may affect the internal validity of the study. However, we segregated analysis based on the method of data collection and the difference was not significant.

## Conclusions

COVID-19 vaccine rejection is low; however, continuous health education and social support is necessary to maintain the high acceptance rates. Time and residency have no significant effect on vaccine acceptance. However, the country-level case fatality and the officially reported number of cases were significant predictors of COVID-19 vaccine acceptance. That’s why encouraging the health authorities to accurately follow & announce case fatalities could be a major contributing factor to increasing vaccine acceptance. We believe that this study will demonstrate public hesitancy and help further development of motivational interview sessions and community-based education tailored according to the population education and individual concerns (73).Although, the main predictor for covid 19 vaccine acceptance or rejection is reporting transparency statement, there are poor transparency of documented information that guide the public decision regarding the vaccine acceptance.Global Health care authorities must report and announce for all transparency international freedom of information act templates (FOIA) to the public for requiring vaccine and providing accurate information regarding all types of vaccines.(74)

## Data Availability

systematic review and meta-analysis

## Abbreviations

AESI: Adverse Events of Special Interest
CAWI: Computer Assisted Web Interviews
CI: Confidence Interval
COVID-19: Coronavirus Disease
F: Female
FDA: Food and Drug Administration
HCWs: Health Care Workers
M: Male
mRNA: Messenger Ribonucleic Acid
n: Number
NA: Not Available
PRISMA: Preferred Reporting Items of Systematic Review and Meta-analysis
QA: Quality Assessment
RRR: Relative Risk Ratio
SARS-CoV-2: Severe Acute Respiratory Syndrome Coronavirus 2
SD: Standard Deviation
SMS: Short Message Service
UK: United Kingdom
US: United States
VAERS: Vaccine Adverse Event Reporting System
VPD: Vaccine Preventable Diseases
WHO: World Health Organization

